# Comparison of the urinary proteome in depressed patients before and after modified electroconvulsive therapy

**DOI:** 10.1101/2023.05.24.23290254

**Authors:** Shuxuan Tang, Tong Guo, Sha Sha, Jing Wei, Yuqing Liu, Jian Yang, Gang Wang, Youhe Gao

## Abstract

Modified electroconvulsive therapy (MECT) is an effective treatment for mood and psychiatric disorders and provides rapid and significant improvements in severe symptoms of several mental health conditions. We attempted to compare the urinary proteome of each depressive patient before and after electroconvulsive therapy. The common biological processes enriched by differential proteins through GO analysis with most patients mainly included cell adhesion (9/9 patients), immune response (7/9), axon guidance (7/9), and oxidation stress (7/9). Moreover, the common biological pathways identified by Ingenuity Pathway Analysis software showed that acute phase response signaling (7/9 patients), NRF2-mediated oxidative stress response (7/9), synaptogenesis signaling pathway (5/9), and ephrin B signaling (5/9) were all upregulated among each patient and were involved in promoting synaptic plasticity and neuroplasticity. Common biological processes and pathways in urine were reported to be related to the mechanism of MECT in the treatment of mental illnesses. In addition, the common differential proteins CSPG4, CBG, APP, NCAM1, and ARSA were suspected to be related to memory loss, memory damage, and memory formation, which might be the effects of MECT.

## 1 Introduction

Major depressive disorder (MDD) is a common psychiatric disorder with a lifetime prevalence of 15-17% (1). Modified electroconvulsive therapy (MECT) has been used in patients for more than 75 years and is considered the most effective treatment for major depressive disorder (2). In the short term, ECT resulted in a remission rate of 70-90% (3). Most MDD patients undergoing MECT require medical maintenance to reduce relapse rates (4).

The main therapeutic effect of ECT is seizures induced by electrical stimulation. The basis and molecular mechanisms underlying the therapeutic effects of MECT are poorly understood. Animal experiments have shown that ECT stimulates the stabilization of cells and synaptic plasticity in the hippocampus, and ECT induces neurogenesis and stimulates neuronal proliferation (5).

Electroconvulsive shock increases glial cell proliferation in the frontal cortex (6). However, MECT caused generalized memory impairment as a side effect, and proteome analysis of the effects of ECS on the brain has been studied in whole brain tissues (7). However, urinary proteome analysis of MECT is lacking.

As a filtrate of the blood, urine bears no need or mechanism to be stable. Urine is the place where most of the waste in blood is dumped, and thus, it tolerates changes to a much higher degree (8). Studies of mental diseases in urine include Parkinson’s disease (9), autism (10) and Alzheimer’s disease (11), and the physiological and pathological changes of these diseases have been detected in urine. We tried to use LC‒MS/MS to explore the mechanism of MECT through the urine proteome. Samples of 9 MDD patients were collected before (BT=9) and after MECT (AT =9). Due to the large heterogeneity of depression and the different types of drugs taken by patients, the individual variability of the sample was large. To eliminate the influence of confounding factors, the self-control comparison method was adopted; that is, the differential proteins were obtained by comparing each patient before and after MECT. Common differential proteins and biological pathways shared by most patients were considered the most likely effects of MECT and were further analyzed. The general workflow is shown in Figure 1.

**Fig. 1.**
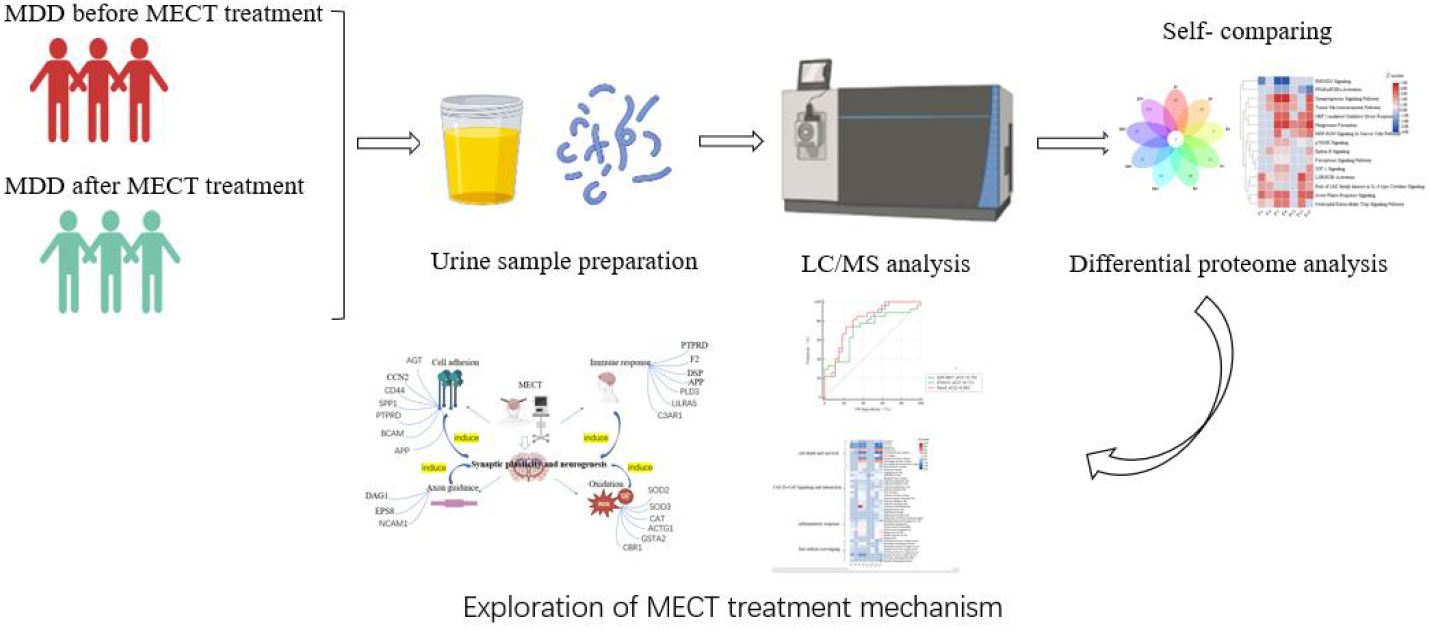
Workflow of this study

## 2. Materials and methods

### 2.1 Participants and assessment

Patients were recruited from the Depression Treatment Center, Beijing Anding Hospital Affiliated to Capital Medical University. Participants signed the informed consent form. Clinicians were asked to screen participants according to depression criteria. The study was approved by the STI2030-Major project (2021ZD0200600) admitted by the Anding Hospital Ethics Review Committee. Participants were included if they (1) met the criteria for DSM-5 severe depression disorder. (2) For the first time or recurrence with a Hamilton Depression Scale (HAMD) score ≥ 17.

(3) Through the collection of clinical medical history and the description of family members, the symptoms of patients’ observations after admission conform to the patient’s MECT indications: severe depression, obvious self-blame, or suicide self-injury concept. (4) Participants signed informed consent.

Subjects commenced ECT with a 0.5 millisecond (ms) pulse width. After 6-8 MECT sessions, the patient was treated for clinical evaluation of symptom improvement. Patient symptoms were assessed between pre- and post-MECT by measuring (1) the Hamilton Anxiety Scale (HAMA) score reduction rate, which was 25%-49%. (2) The Hamilton Depression Scale (HAMD) index is less than 17. Patients were considered to have improved clinical symptoms. Depression symptoms improved in these patients after MECT. A total of 18 samples were enrolled, and they were divided into two groups: before MECT (BT N=9) and after MECT (N=9). The urine samples were stored at -80 °C as soon as possible.

### 2.2 Sample preparation

A urine sample of 3-4 ml was centrifuged at a low speed of 3000*g for 10 min, and cell fragments were removed. The supernatant was denatured by the working concentration of 20 mm dithiothreitol at 95 ℃ for 10 minutes and restored to room temperature. Then, they were alkylated in a working concentration of 50 mM iodoacetamide in the dark for 45 min. The urine samples were precipitated overnight using 6 times the volume of ethanol at -20 °C for 40 min and centrifuged at 14000*g for 30 min at 4 °C, and the pellets were resuspended and dissolved in 20 mM Tris buffer. The protein concentration was quantified by a Bradford kit. The 10 kD ultracentrifugation filters were repeatedly washed three times with 20 mM Tris buffer. The 100-µg protein sample placed on a 10 kDa ultrafiltration membrane was centrifuged (14000*g) at 4 °C for 30 min. The protein on the ultrafiltration membrane was repeatedly pipetted with Tris and then centrifuged. The process was repeated 3-4 times to ensure that there was no liquid in the sleeve for the last time. The ultrafiltration tubes were placed in another new casing, and trypsin (1:50) was added to these samples and then incubated at 37 °C overnight. After digestion, the digested peptides were eluted from 10-kD filters and desalted using HLB (Waters, Milford, MA, USA).

### 2.3 Peptide fractionation

The peptides were diluted to 0.5 µg/µl, and a total of 2 µg of each sample was mixed. The pooled peptide was separated on high pH reversed-phase spin columns (Thermo Scientific, 84868). Eluents with different concentrations of acetonitrile (5, 7.5, 10, 12.5, 15, 17.5, 20 and 50%) were loaded on reversed-phase columns and centrifuged for 2 min per fraction. A total of 10 fractions were lyophilized and dissolved in 20 µl of 0.1% formic acid.

### 2.4 Liquid chromatography-tandem mass spectrometry analysis

The Orbitrap Fusion Lumos (Thermo Scientific) coupled with an Easy 1000 was used for LC‒MS/MS analysis. The digested peptides were separated on a C18 high resolution column (Omitech, 75 μm × 150 mm). Peptides were eluted with a gradient of 4%–35% buffer B (0.1% formic acid in 80% acetonitrile, flow rate at 0.3 µl/min) for 90 min.

Two microliters of each sample were added to a pooled mixed sample used as quality control (QC). Every 10 samples were inserted into one QC sample to monitor the reproducibility of the LC‒MS/MS. To generate the spectral library, fractions were analyzed in data-dependent acquisition (DDA) mode. The parameters were set as follows: the scan range was set at 350-1550 m/z; the MS resolution was 120000; the maximum injection time was 50 ms; the auto gain control (AGC) target was 4.0e5; and the cycle time was 3 s. MS/MS resolution was performed at 30000; the collision energy was set at 32% (HCD); the AGC target was 5.0e^4^; the maximum injection time was 45 ms; and the exclusion duration was 30 s.

Each sample and the QC samples were analyzed in data-independent acquisition (DIA) mode. For DIA MS acquisition, a variable 36 isolation window was generated. The specific window lists were constructed based on fractions acquired by DDA mode. The full scan was set at a resolution of 60000 over the m/z range of 350-1500; the AGC target was 1.0e^6^; and the maximum injection time was 50 ms. DIA scans with a resolution of 30000. The AGC target was set to 1.0e^6^; the HCD collision energy was 30%, and the maximal injection time was 100 ms.

### 2.5 Spectral Library Generation

To generate a spectral library, the fraction DDA data were processed by Proteome Discoverer 2.1 (Thermo Scientific, USA) software searched against the human UniProt database attached to the irt sequence. A maximum of two missed cleavages for trypsin was allowed, cysteine carbamidomethylation was set as a fixed modification, and oxidation (M) was set as a variable modification. The parent and fragment ion mass tolerances were set to 10 ppm and 0.02 Da, respectively. The applied false discovery rate (FDR) cutoff was set to less than 1%. Then, the results were imported to Spectronaut Pulsar (Biognosys AG, Switzerland) to generate the spectral library. Next, QC and samples and the fraction libraries were used to one urine spectral library with default settings of Spectronaut Pulsar. The peptide retention time was calibrated according to the iRT data. Peptide intensity was calculated by summing the peak areas of their respective fragment ions for MS2. Cross-run normalization was enabled to correct for systematic variance in the LC‒MS performance, and a local normalization strategy was used. All results were filtered by a Q value cutoff of 0.01 (corresponding to an FDR of 1%).

### 2.6 Data analysis

For quantified proteome data, proteins with at least two unique peptides were allowed. QC was used as technical replicates to monitor instrument stability. Proteins with QC<0.3 were retained, and missing values of QC and sample were filled using the K-NN method on the Wukong website (https://www.omicsolution.org/wkomics/main/). The remaining proteins in the two groups of patients before and after MECT with missing values >0.5 were removed, and the remaining proteins were finally used as the final data to analyze the differential proteins. The differential proteins were screened with the following criteria: proteins with at least two unique peptides were allowed; fold change ≥ 1.5 or ≤ 0.65; comparison of individual urine proteome before and after MECT by t test, and p < 0.05 were identified as statistically significant. The P values were adjusted by the Benjamini and Hochberg method on the Wukong website (https://www.omicsolution.org/wkomics/main/). The results are presented as the mean ± SD.

### 2.7 Bioinformatics analysis

Differential proteins were used for biological process through GO analysis on the David website and for canonical pathways by Ingenuity Pathway Analysis software (Ingenuity Systems, Mountain View, CA). The proteins were mapped to disease and function categories by IPA software ranked by P value. PCA was performed by SIMICA 14.1 software. Heatmaps were made by Tbtools software. Pearson’s correlation coefficient was determined through the Wukong website. Histograms and scatterplots were generated with GraphPad Prism 8. The biomedical sketch picture comes from the website https://app.biorender.com/user/signup

## 3. Results and discussion

### 3.1 Demographic characteristics of participants

In this study, 18 samples of major depression disorder patients were recruited, including 9 before treatment patients and 9 after treatment patients. The detailed clinical characteristics of the participants are shown in Table 1. The recruited patients met the clinical criteria for depression and met the indications for MECT treatment. After MECT treatment, the depressive symptoms of 9 patients were significantly improved according to the HAMD scale. Urine samples were retained within 24 hours after completion of MECT. The urine samples were analyzed using the DIA method. Because of the large heterogeneity of depression, confounding factors such as the type of medication, age, and pathogenesis of depression among individual patients lead to large interindividual differences, and before and after MECT of urinary proteome comparisons for individual patients were adopted. The differential proteins and enriched biological functions of each patient were used for setting operation, that is, to find the functions or proteins shared by most patients to further explore the mechanism of MECT treatment. The workflow is shown in Fig. 1.

**Table 1.**
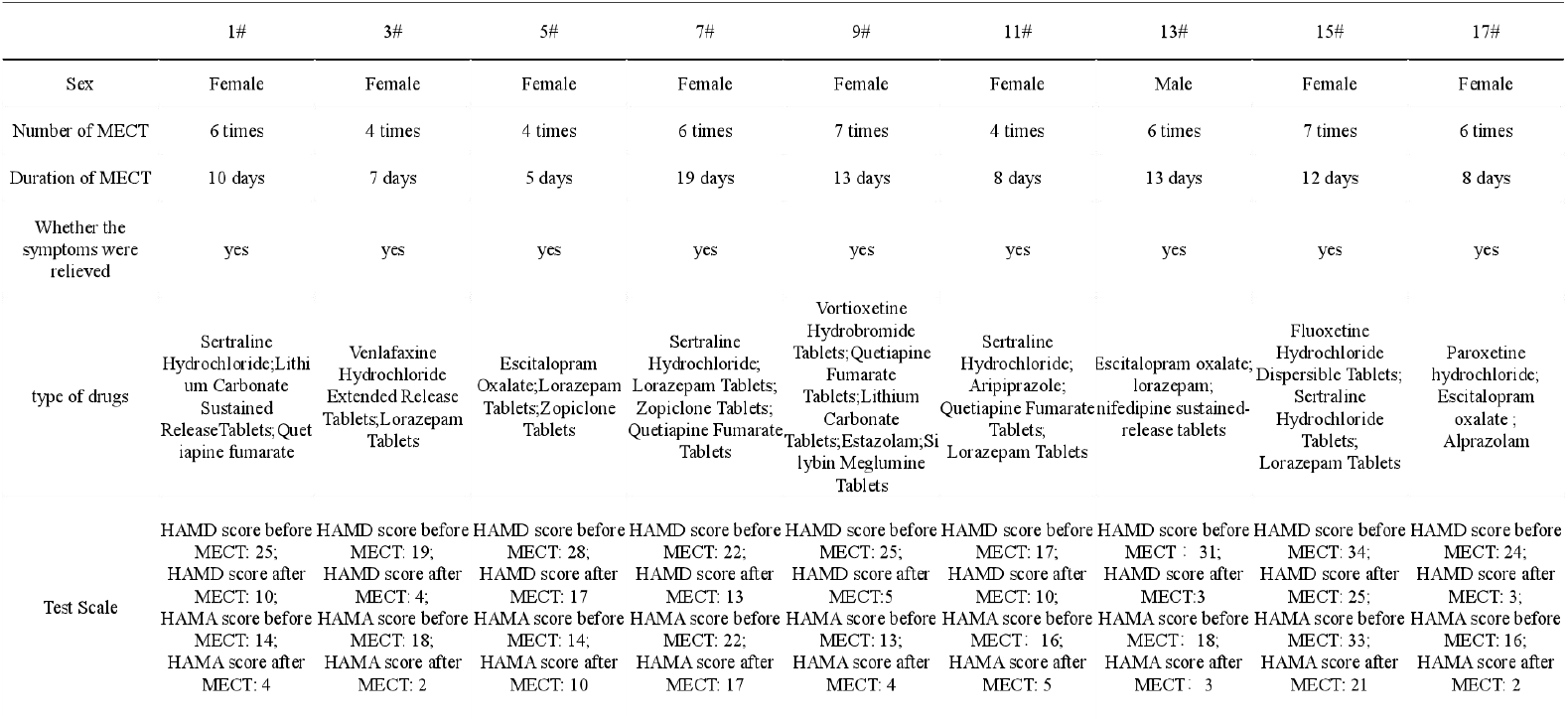
Demographics of participants in the study

### 3.2 The urinary proteome profile of BT and AT patients

A total of 18 urine samples from MDD patients before and after treatment were analyzed using the DIA method. Quality control (QC) was a pool consisting of each sample aliquot. QC was used as a quality control to monitor the stability of the instrument, and a QC sample was inserted every 10 samples. Each sample was subjected to mass spectrometry in three technical replicates. The 61 samples were used to construct a library with a total of 3108 proteins, and a total of 2490 proteins with an FDR <1.0% and at least 2 unique peptides were identified. Technical variation of this analysis was evaluated by calculating the coefficient of variance (CV) of QC replicates. The median CV of QC technical replicates was 0.17, and over 73% quantile of QC technical replicates was less than 0.3. The protein groups identified for each sample within the group are shown in Fig. 2B, and each patient in the BT treatment and AT treatment groups was 2342±498.4 and 2484± 258.8. The median BT interindividual CV and AT interindividual CV were 1.225 and 0.7, respectively.

**Fig. 2.**
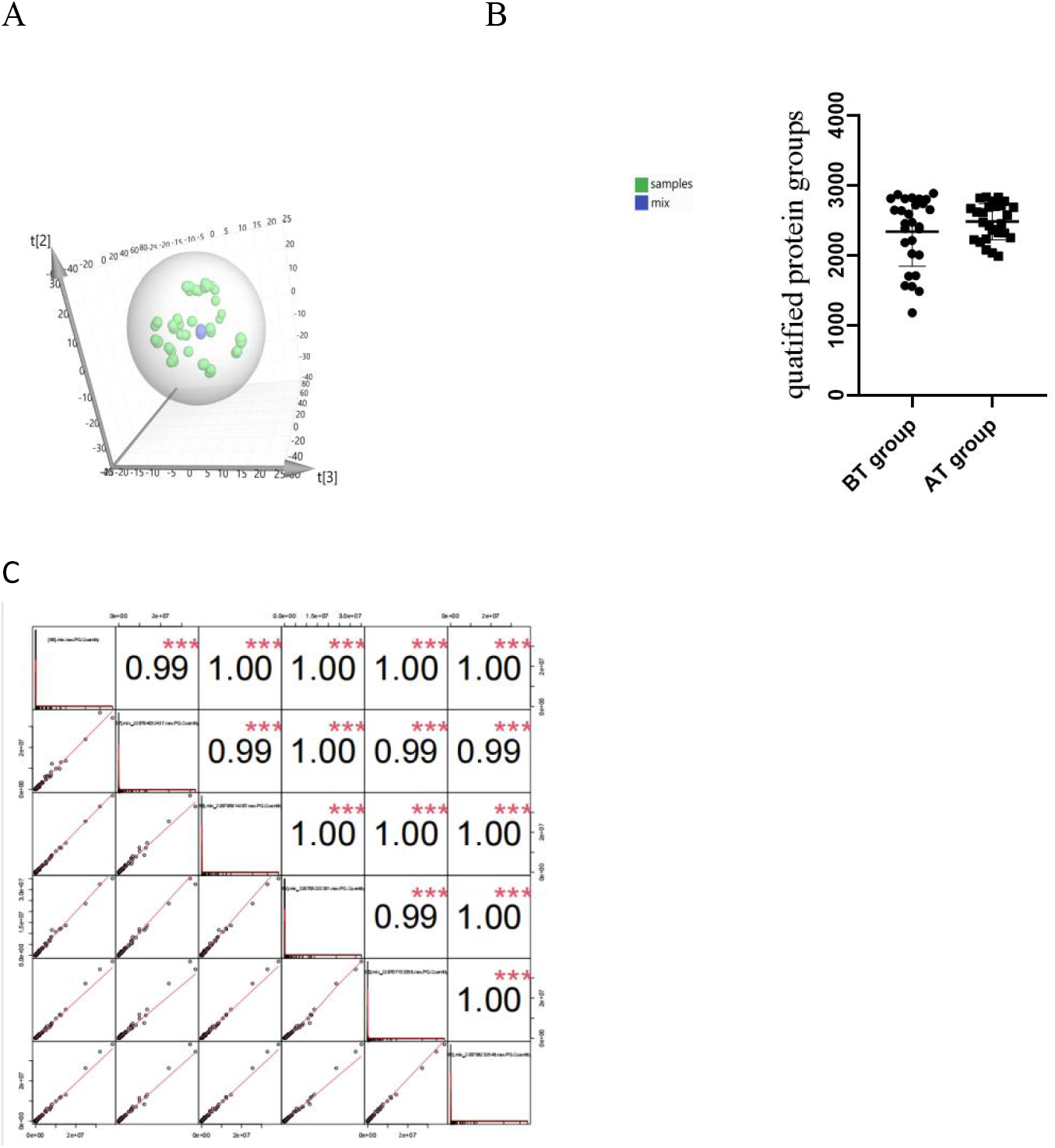
Urinary proteome profile. (A) PCA plots of mixed and patient samples. (B) Number of proteins identified per patient within the BT and AT groups. Quantities are presented as the mean ± SD. (C) Pearson coefficient correlation of QC

The PCA cluster analysis showed that QC samples were clustered together (Fig. 2A). Pearson’s correlation coefficients were approximately 1 among 6 QC replicates (Fig. 2C) and showed good technical reproducibility. Proteins with CV values of QC less than 0.3 were retained, leaving 1594 proteins as the final dataset for subsequent analysis after missing values were filled(supplematary table1).

### 3.3 Urinary proteome changes before and after MECT

The confounding factors of MDD, such as taking drugs and age, have a greater impact on the urinary proteome. To observe the changes generated by MECT in urine more obviously, the differential proteins were obtained by comparing each patient before and after treatment. The number of differential proteins for 9 patients was 342, 585, 416, 142, 564, 215, 100, 384, and 784, respectively (Fig. 3A). The differential proteins for each patient were submitted to the canonical pathway and biological process by IPA and DAVID. The number of biological processes and pathways enriched in 9 patients were 153, 249, 187, 36, 273, 113, 46, 215, 327 and 86, 44, 183, 18, 153, 44, 61, 118, 277, respectively. (Supplementary Table 2 and Table 3). There were no common proteins or canonical pathways for 9 patients, and 3, 8, and 33 common differential proteins were counted in 8/9, 7/9, and 6/9 patients, respectively (Table 2). The number of common biological processes by GO analysis for 9 patients was 3, and 7 and 18 common biological processes were counted in 8/9 and 7/9 patients, respectively (Table 2). The number of common canonical pathways by IPA for 7/9, 6/9 patients was 5, 16, and 32 respectively (Table 2). The log_2_FC value of the common differential protein (percent of patient number >60%) before and after MECT showed that the correlation was low among 9 patients, suggesting that the physiological results of electrotherapy improving depressive symptoms were different in 9 patients (Fig. 3B).

**Table 2.** Common differentially expressed proteins, biological processes, and canonical pathways.

|  |  |
| --- | --- |
| Differential proteins enriched by number of patients | Number of common protein |
| 8/9 | 8 |
| 7/9 | 3 |
| 6/9 | 33 |
| Common biological process enriched by number of patients | Number of biological process |
| 9/9 | 3 |
| 8/9 | 7 |
| 7/9 | 18 |
| 6/9 | 9 |
| Common canonical pathways enriched by number of patients | Number of canonical pathways |
| 7/9 | 5 |
| 6/9 | 16 |
| 5/9 | 32 |

**Table 3.**
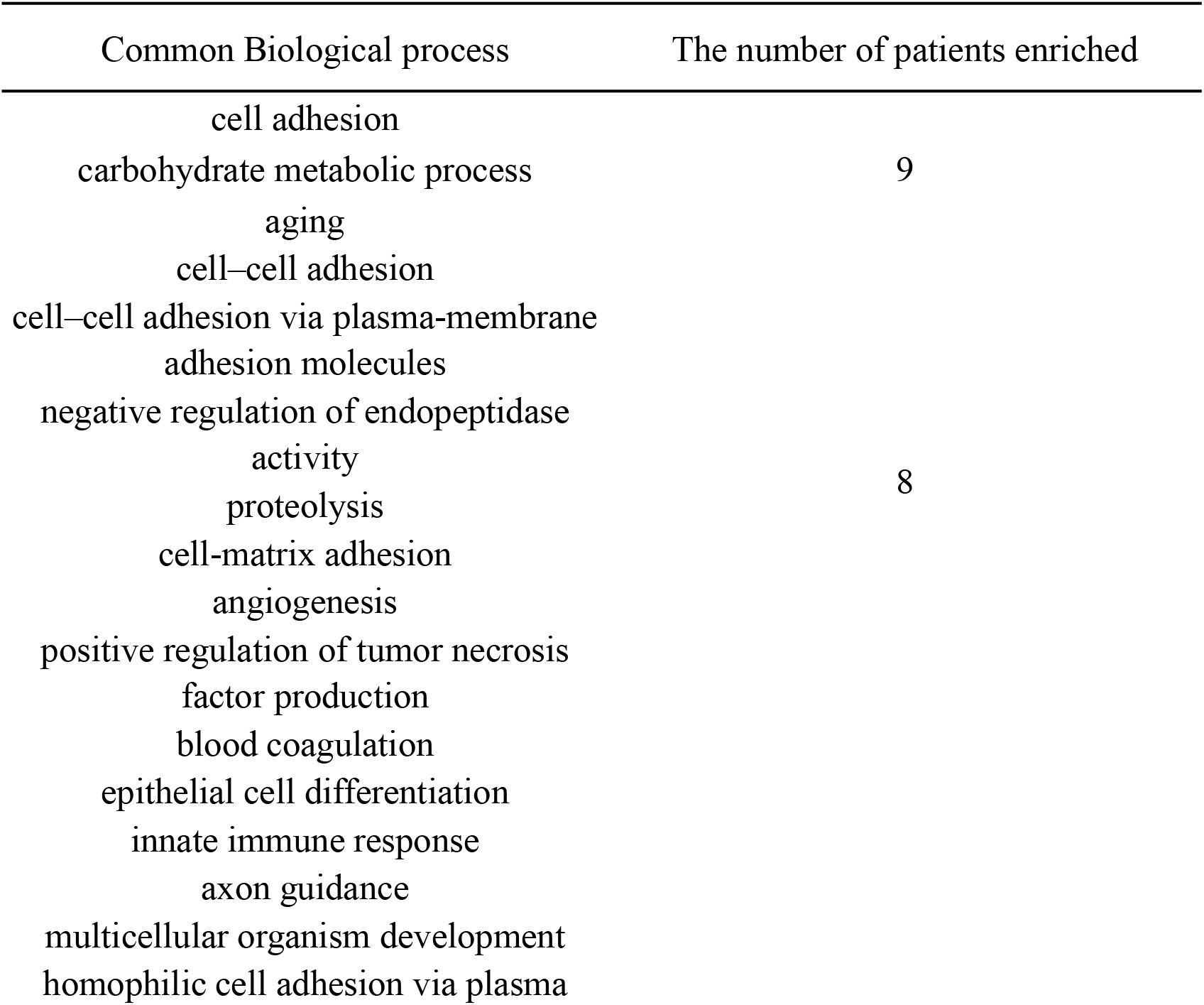
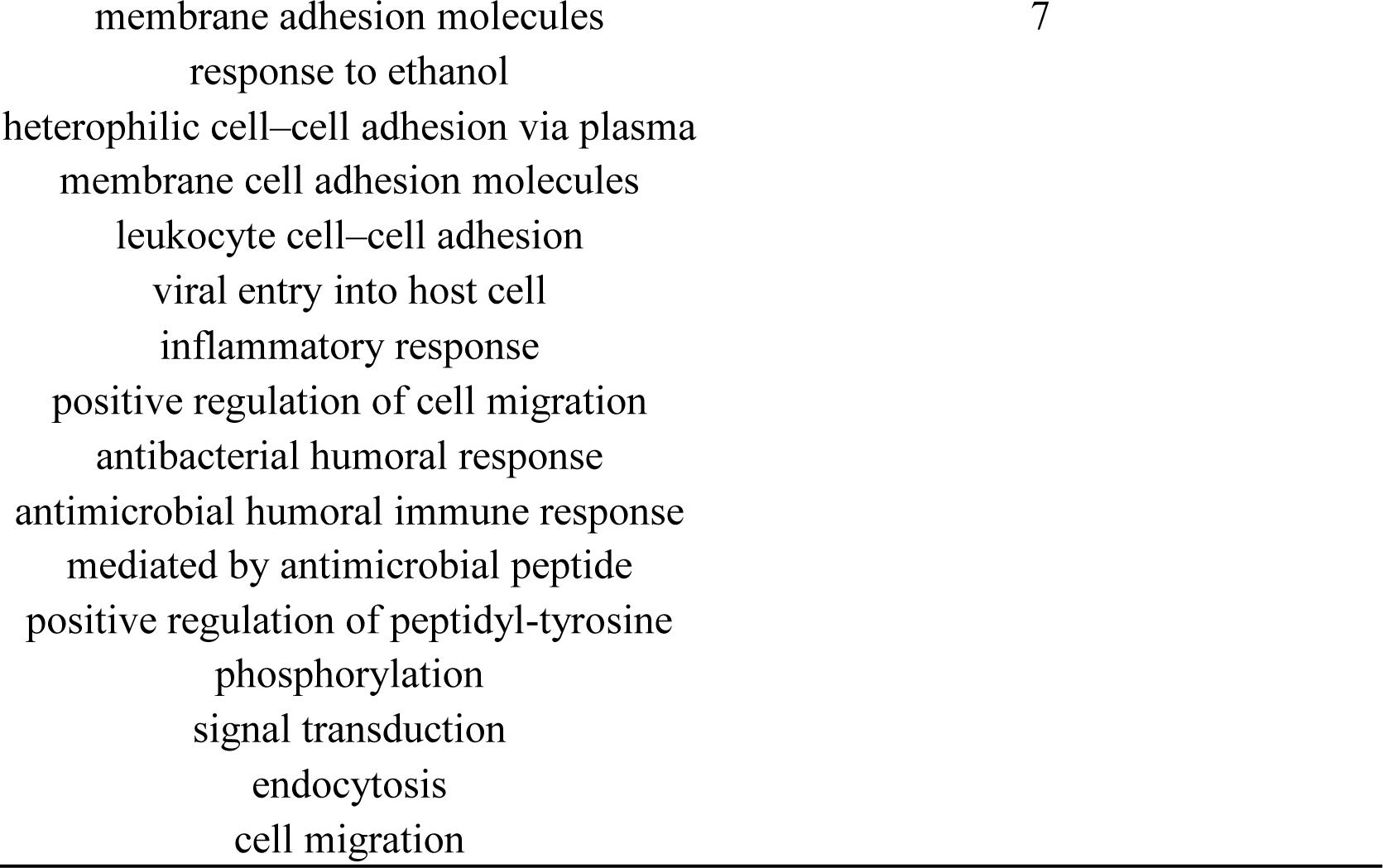
Common biological process enriched by most patients

**Fig. 3.**
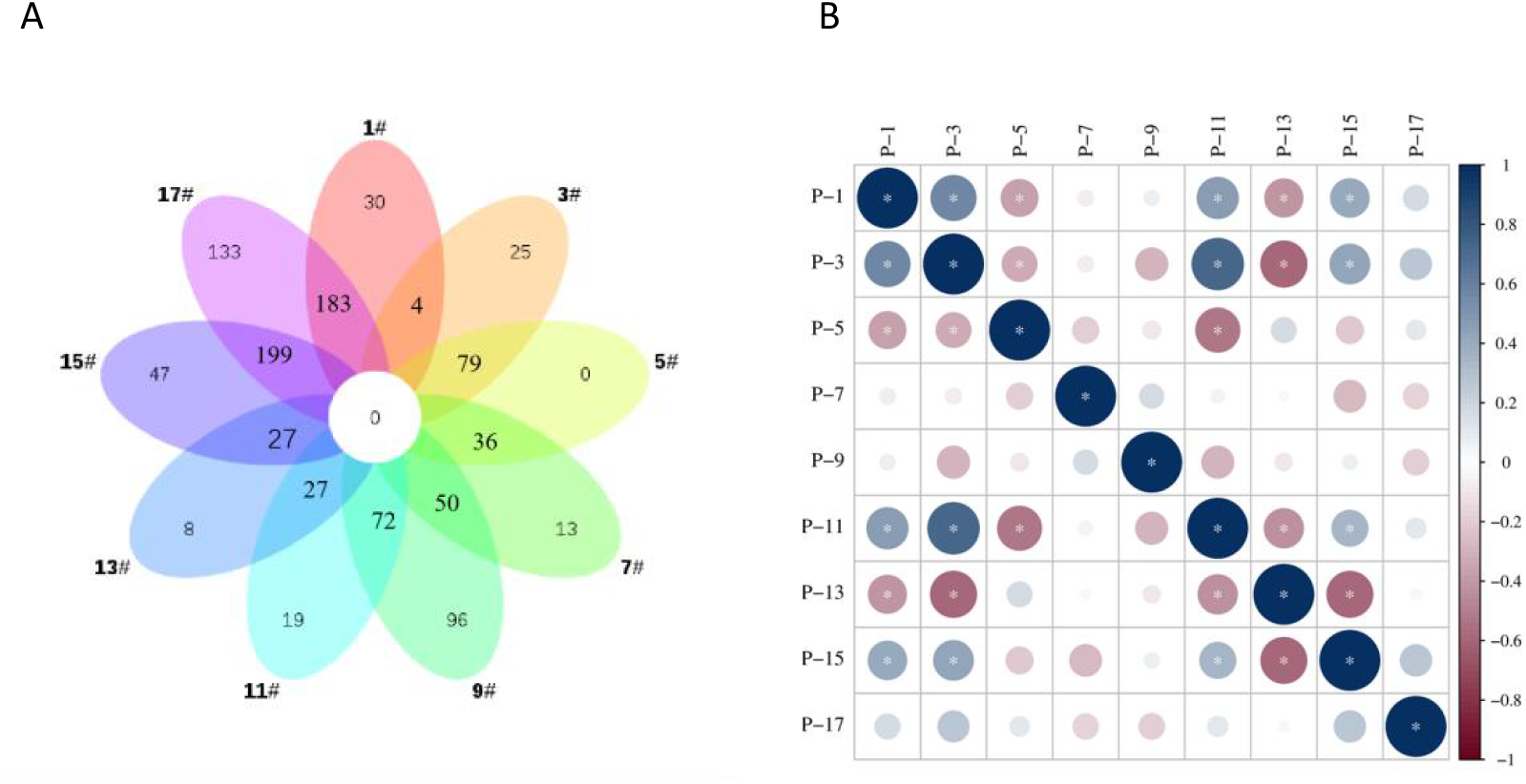
Venn diagram of the differentially expressed proteins

### 3.4 Functional annotation of differential proteins

The biological processes involved in the common differentially expressed proteins (N>60%) are shown in Fig. 4. According to the significance, the top pathways were cell adhesion, positive regulation of cell activation, inflammatory factor response pathway, and astrocyte activation process (P value < 0.05). Specific neuroendocrine-immune interactions coordinated by astrocytes contribute to depressive symptoms; therefore, standard treatment is not effective (12). Targeted reduction of astrocyte-mediated neuroinflammation has been reported to alleviate depression-like behaviors (13). The differential proteins of each person were submitted to the biological functions. The biological processes and canonical pathways enriched for each patient were statistically analyzed. The biological processes and canonical pathways top distributed in most patients were analyzed (Supplementary table 4A and table 4B). The common processes in all patients (N=9) were cell adhesion, carbohydrate metabolic process, and aging; those in 8 patients (N=8/9) were cell‒cell adhesion, cell‒cell adhesion via plasma-membrane adhesion molecules, negative regulation of endopeptidase activity, proteolysis, cell-matrix adhesion angiogenesis, and positive regulation of tumor necrosis factor production; and those in 7 patients (N=7/9) were innate immune response, axon guidance, multicellular organism development, homophilic cell adhesion via plasma membrane adhesion molecules (Table 3).

**Table 4.**
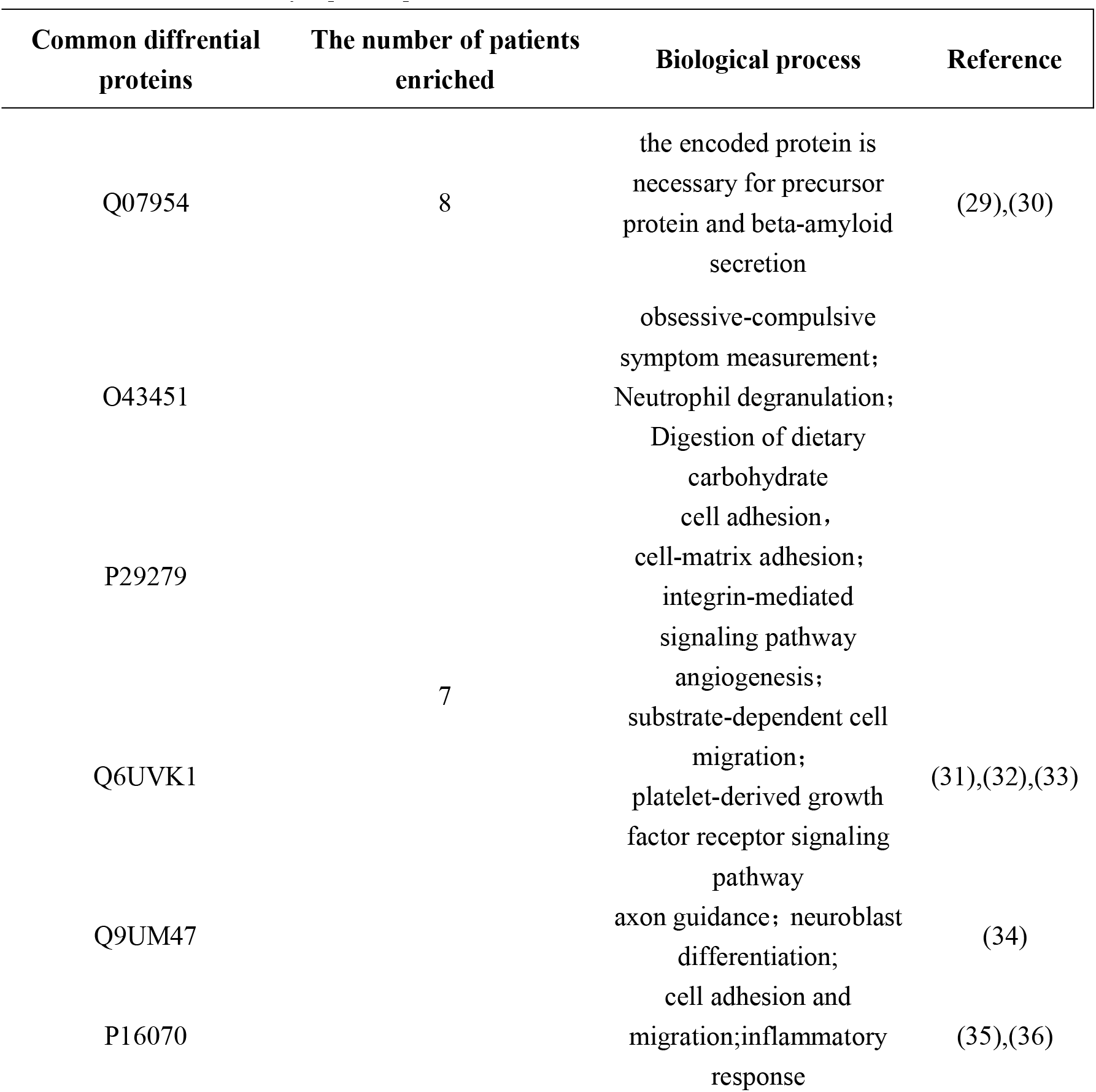
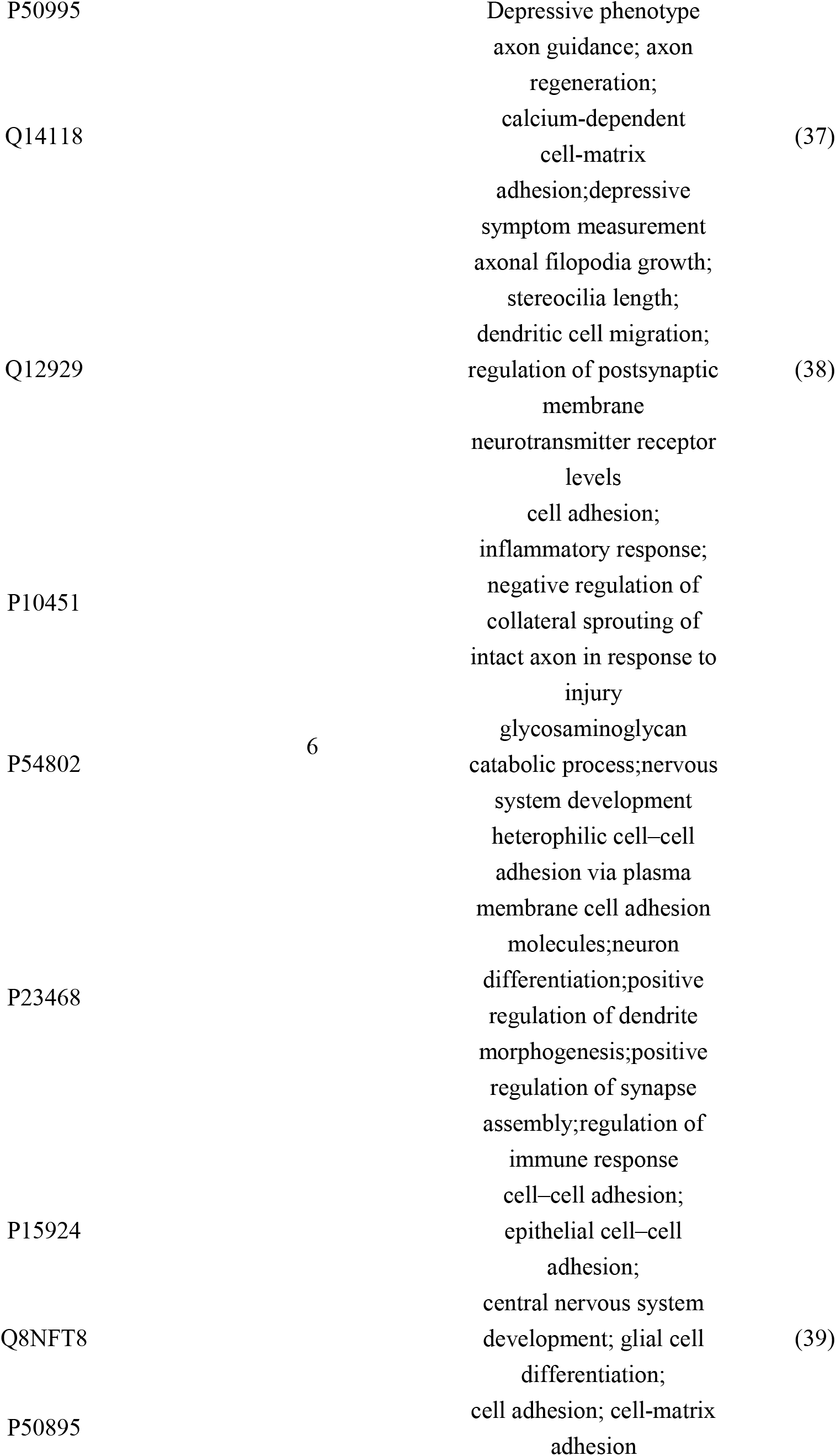
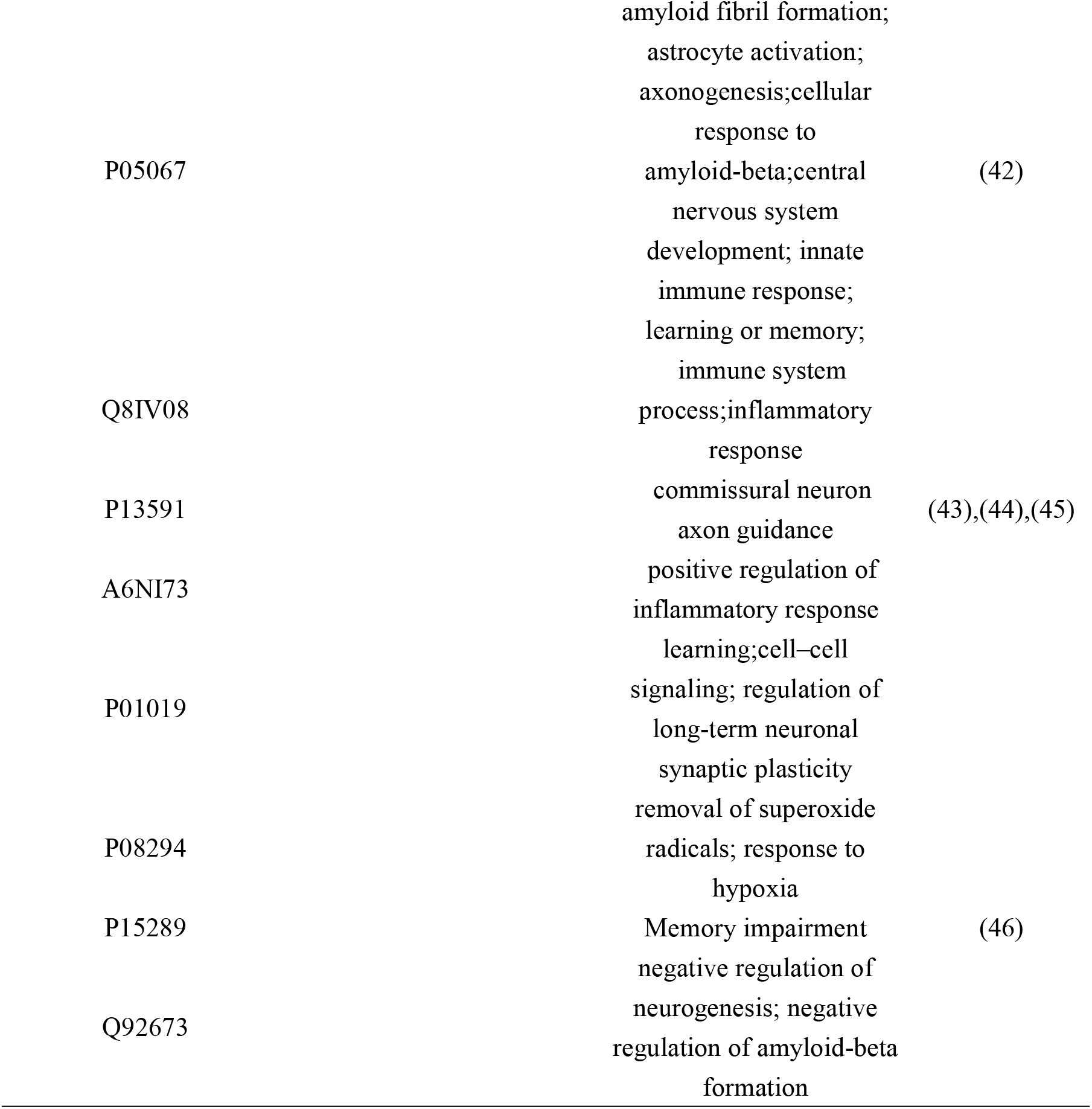
Common differentially expressed protein MECT-related annotation

**Fig. 4.**
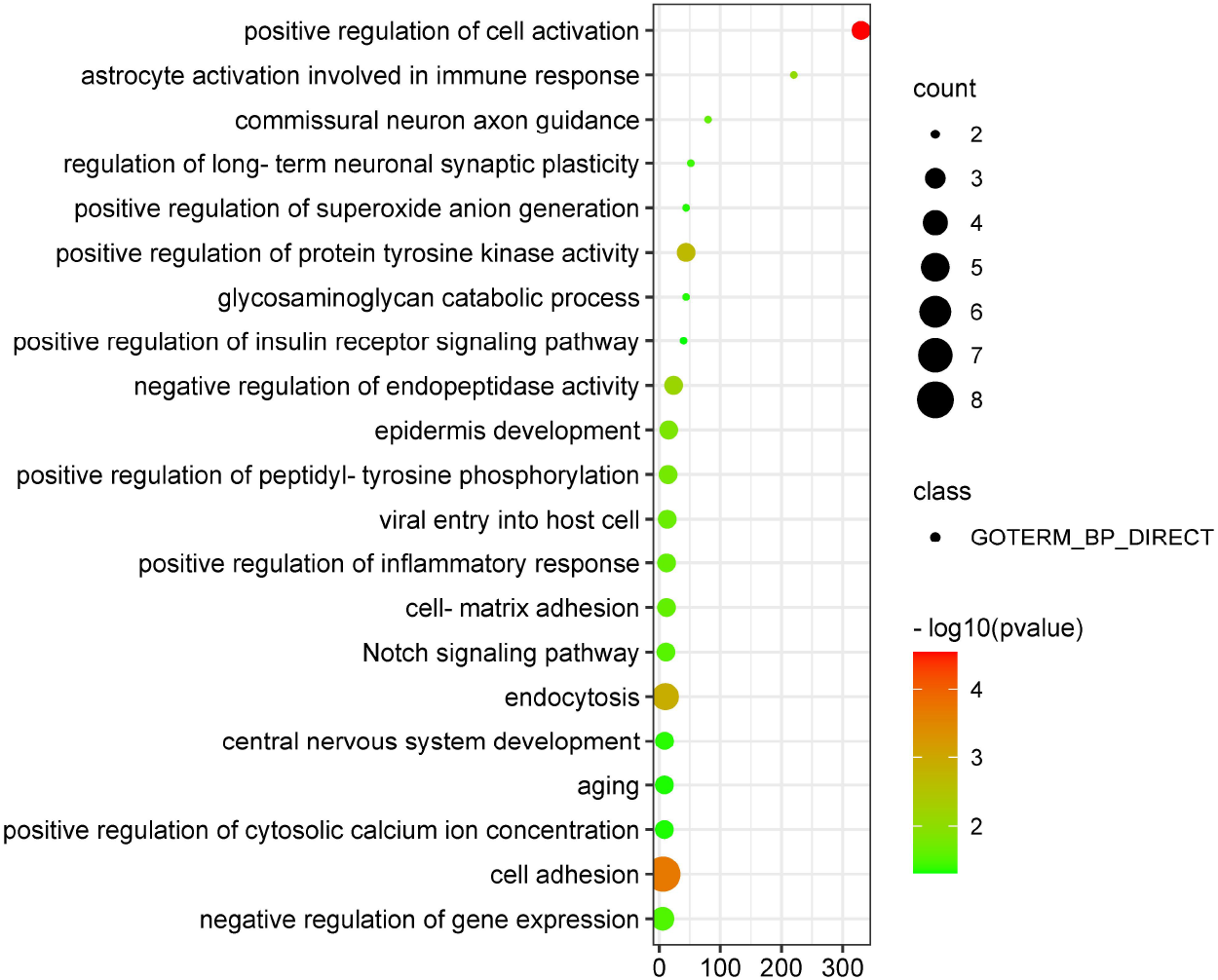
Biological processes enriched by common differential proteins among patients.

The biological processes related to cell adhesion were obviously detected in 9 patients. The hippocampus is a site of active neurogenesis and neuroplasticity, and ECT may induce neurogenesis, synaptogenesis, and glial proliferation (12). Adhesion molecules are involved in target recognition and stabilization during synapse formation. They are required for axon development from initially inducing outgrowth to guiding targets (13). Emerging mechanisms by which adhesion factor Ig-CAMs promote synaptic plasticity include modulation of the activity of NMDA receptors and L-type Ca2þ channels, signaling through the mitogen-activated protein kinase p38(14). The biological processes of tumor necrosis factor (TNF) production were detected in 8 patients. TNF-α regulates synaptic scale by promoting the insertion of AMPARs in the synaptic plasma membrane. TNF-α also regulates glial cell transmission by increasing astrocytic calcium levels and activates neuronal NMDARs at synapses on dentate gyrus granule cells, resulting in increased excitatory synaptic activity and potentiation of GC synapses (15). The innate immune response and axon guidance were common processes in 7 patients. Single ECT induces a transient (15–30 min) increase in the expression of pro-inflammatory cytokines, such as TNF-α, IL-1β and IL-6(16). Specifically, inflammation was associated with changes in the hypothalamic axis and neurogenesis. After MECT treatment, inflammatory factors such as IL-6 levels were significantly decreased, indicating that depression symptoms improved (16). Axon guide proteins guide axon growth during development and control the structural plasticity of synaptic connections (17). The differential urinary proteome before and after MECT induced related neurogenesis and synaptic plasticity pathways consistent with the mechanism of MECT treatment.

The statistical results of the canonical pathways by IPA software are shown in Supplementary Table 4. Common canonical pathways enriched by most patients (N=7/9, 6/9, 5/9) were LXR/RXR activation, acute phase response signaling, NRF2-mediated oxidative stress response, ferroptosis signaling pathway, role of JAK family kinases in IL-6-type cytokine signaling, synaptogenesis signaling pathway, neutrophil extracellular trap signaling pathway, p70S6K signaling, tumor microenvironment pathway, ephrin B signaling signaling, MSP-RON signaling IGF-1 signaling, and phagosome formation, which were upregulated in each patient with enrichment. RHOGDI signaling and PPARα/RXRα activation were downregulated in 7 patients (Fig. 5A).

Superoxide radical degradation was enriched in patients according to IPA software analysis.

Superoxide radical degradation is a reactive oxygen species (ROS) that is involved in several regions of the synaptic plasticity process as a second messenger. Nervous system ROS production is required for hippocampal long-term potentiation (LTP), a form of synaptic enhancement involved in learning and memory in certain types of mammals (18). Nrf2, as a master regulator of metabolism, protein quality control, and antioxidant defense, is involved in neurogenesis. Nrf2 affects the survival, differentiation, and neurogenesis of neural stem cells via ROS (19).

Ferroptosis, a novel form of cell death characterized by mitochondrial damage, oxidative stress, and mitochondrial dysfunction, affects specific types of synaptic plasticity in the spinal cord (20).

The role of JAK family kinases in IL-6-type cytokine signaling was enriched in the urine of patients before and after MECT. Elevated IL-6 levels may indicate greater neuroplasticity capacity through enhanced BDNF secretion, and IL-6, TNF-α, and IL-1-β regulate cognitive function through synaptic plasticity, which is critical for memory formation and retention in the hippocampus. Aryl hydrocarbon receptor signaling was a significant pathway detected in 5 patients. The aryl hydrocarbon receptor (AhR) acts as a regulator of ependymal glial cell differentiation into postmitotic neurons. AhR signaling is a key regulator of the restoration of neurogenesis in the zebrafish brain (21). p70S6 kinase may be important for maintaining long-term potentiation in the hippocampus by regulating actin dynamics, and p70S6K plays an important role in synaptic plasticity and higher brain functions such as learning and memory (22). Ephrin-B2 astrocytes promote neuronal differentiation of adult NSCs through the dynamin signaling pathway (23). Ephrin-B signaling performs a remarkable series of functions throughout the synapse life cycle. EphB is required for dendritic filopodia movement during synaptogenesis, which appears to allow filopodia to ‘interview’ potential synaptic partners in a contact-dependent manner (24). Insulin-like growth factor-1 (IGF-1) is thought to promote neurogenesis and survival (25). IGF-I affects the proliferation and differentiation of neural stem cells into neurons and glial cells and neuronal maturation, including synapse formation (26). The phagosomal pathway plays an important role in tissue homeostasis, antigen presentation and autophagy. (27). Autophagy maintains neuronal homeostasis and viability by preventing the accumulation of toxic and pathological intracellular aggregates. Therefore, autophagy plays a multifaceted role during the occurrence of nerves from stem cells to differentiation of nerve cells (28). The above canonical pathways were related to neurogenesis, synaptic plasticity and LTP formation. Patthways are relevant to the relationship between hippocampal neuroplasticity and therapeutic mechanisms promoting neurogenesis and other neuroplastic effects including gliogenesis, angiogenesis, and synaptogenesis(29).

Disease and function analyses showed that cell necrosis and apoptosis, cell death of tumor cell lines, and apoptosis of tumor cell lines were deactivated, and cell survival, cell viability, cell viability of tumor cell lines, cell viability of nervous tissue cell lines and survival of stem cell lines were activated. Cell-to-cell signaling and interaction-related pathways including aggregation of cells, binding of tumor cell lines, adhesion of epithelial cells, adhesion of immune cells, adhesion of blood cells, cell‒cell contact, adhesion of tumor cell lines, phagocytosis of cells and binding of blood cells were activated. Immune response-related processes and free radical scavenging-related metabolism of reactive oxygen species were activated (Fig. 4B). The above results indicated that urine could reflect changes in the nervous system and immune response generated by MECT.

**Fig. 5.**
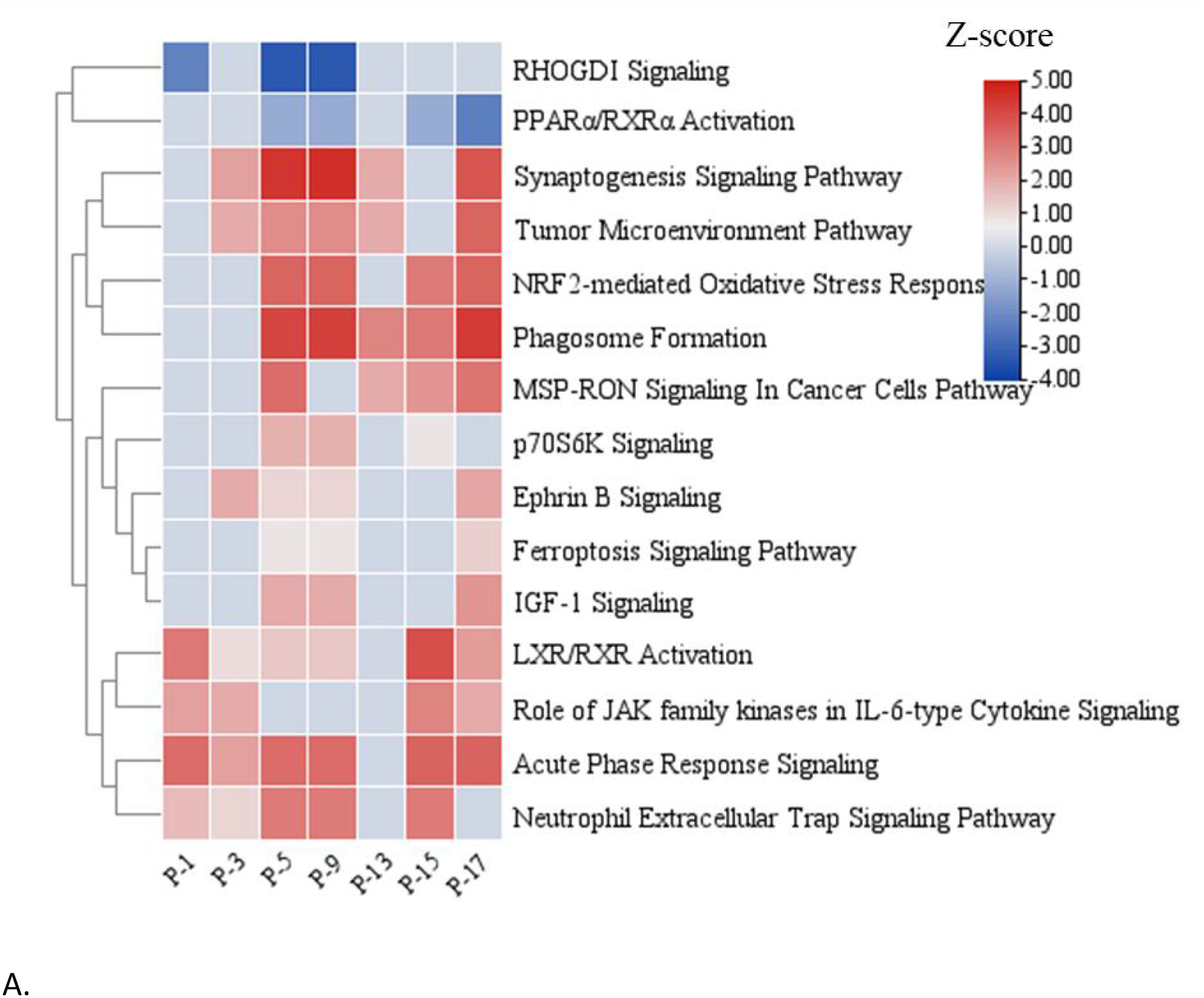
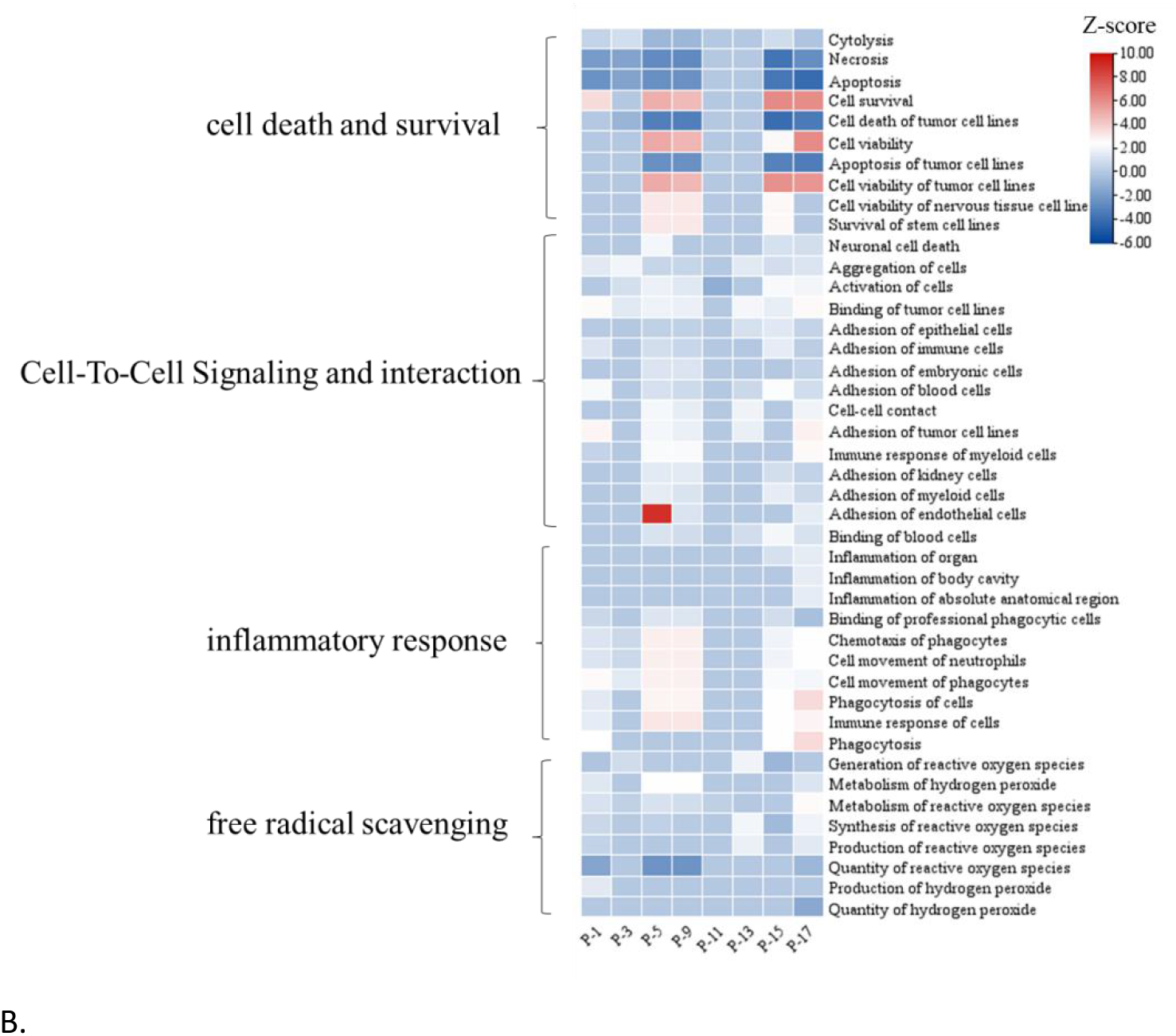
Functional annotation of the differential urinary proteome before and after MECT. The z score was calculated to predict the trend. A z score with a positive value indicates activation, and a negative value indicates inhibition. A. Common top distributed canonical pathway analyses of MECT for 9 patients. B. Common disease and function analyses of MECT for 9 patients

### 3.5 Differential proteins distinguished before and after MECT analysis

Differential proteins were acquired before and after MECT. Self-control was used to exclude the influence of confounding factors to identify proteins induced by the MECT effect. The differential proteins of each patient were all combined for statistical analysis. To detect significant changes by MECT, common differential proteins by most patients were highlighted. The common differential proteins shared by 8/9, 7/9 and 6/9 patients were 8, 3 and 33, respectively (Supplementary Table 5). The AUC curve was used to evaluate the accuracy of differential proteins to distinguish between before and after MECT. The highest AUC values of protein Q9UM47 (NOTCH3) and P10451 (Osteopontin) were 0.781 and 0.713, respectively, and the sensitivity and specificity were above 70%. The AUC area of their combined panel was 0.802, and the sensitivity and specificity were 85.19% and 66.67%, respectively (Fig. 5). The top proteins in terms of AUC value were NOTCH3, osteopontin, desmoplakin, extracellular superoxide dismutase SOD3 and CCN family member 2 (CCN2). The expression differences of these proteins in each patient are shown in Supplementary Fig. 5. The variation in the trend of differential proteins among patients indicates that although each patient’s depressive symptoms improved, the treatment of depression by MECT was individual in the urinary proteome.

**Fig. 5.**
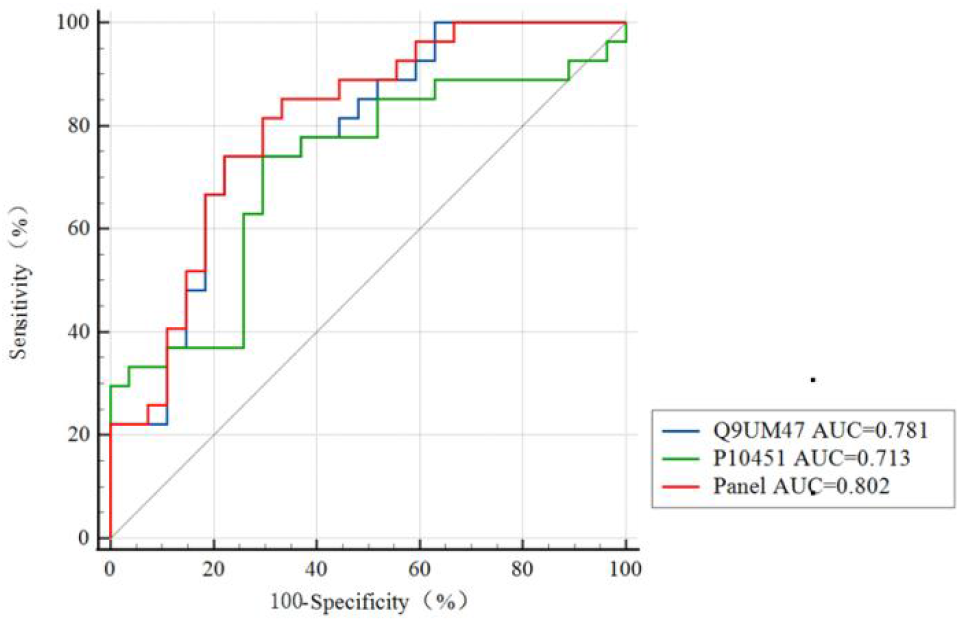
The common differentially expressed proteins induced by MECT were analyzed. Receiver operating characteristic (ROC) curve analysis was performed by Q9UM47, P10451, and the panel (combining Q9UM47 and P10451) to distinguish between before and after MECT. B. The expression levels of common proteins with high AUC values in each patient before and after MECT.

### 3.6 Potential mechanisms associated with MECT reflected in the urinary proteome

To find the changes in the nervous system after MECT in urine. The proteins shared by most patients shown in Table 4 were the most likely proteins related to the mechanism of MECT (Supplementary Table 5).

Q07954, neuronal prow-density lipoprotein receptor-related protein 1 (LRP1) plays an important role in regulating blood‒brain barrier permeability, neuronal growth, and calcium signaling through essential signal transduction pathways in the central nervous system (30). We found that LRP1 expression was positively correlated with depressive-like behaviors (31). P29279 and CCN family member 2 (CCN2) are involved in cell adhesion, cell-matrix adhesion and the integrin-mediated signaling pathway. Q6UVK1 and chondroitin sulfate proteoglycan 4 (CSPG4) provide growth cone guidance cues to the developing central nervous system (CNS) and contribute to the formation of neuronal boundaries, and their presence in the perineural network plays a crucial role in the maturation of synapses (32). CSPGs were hypothesized to maintain neuronal connections and attenuate structural plasticity or rearrangement of synapses in the adult CNS(33). CSPGs are associated with the formation of traumatic memories(34). Q9UM47, Neurogenic locus notch homolog protein 3 (NOTCH3), is involved in axon guidance and neuroblast differentiation. Previous results have established that Notch also regulates connectivity in the nervous system and acts at different levels, including the specification of neuronal identity, division, survival, and migration, as well as axon guidance, morphogenesis of dendritic arbors, and synaptic regulation. increase in touch strength(35). P16070 and CD44 antigen (CD44) play roles in cell‒cell interactions, cell adhesion and migration and the inflammatory response. CD44 adhesion molecule inhibited dendritic tree arborization in vitro and in vivo(36). CD44 affects synaptic excitatory transmission in primary hippocampal neurons while altering dendritic spine shape(37). Q14118, dystroglycan 1 (DAG1) relies on its extracellular scaffold function to regulate the development of multiple axonal tracts in the spinal cord and brain. Dystroglycan acts as a multifunctional regulator that directs axons throughout the nervous system by coordinating multiple ECM proteins, secreted cues, and transmembrane receptors(38). Q12929, Epidermal growth factor receptor kinase substrate 8 (EPS8).

Epidermal growth factor receptor (EGFR) is the target of choice for the treatment of Aβ-induced memory loss. The EGFR pathway can regulate neuronal plasticity by altering intracellular Ca^2+^ concentration or glutamate release in postmitotic neurons(39). Q8NFT8, Delta and Notch-like epidermal growth factor-related receptor (DNER) are expressed in spiral ganglion neurons and appear to be involved in the mechanisms of neuronal polarity and neuroclastogenesis through Notch-dependent signaling pathways (40). P08185, Corticosteroid-binding globulin(CBG) role in hippocampal-dependent memory consolidation in mice. Given the importance of CBG in modulating the effects of glucocorticoids on memory, it is an interesting target for drugs to treat memory-related disorders known to be affected by glucocorticoids, such as posttraumatic stress disorder or phobias (41). CBG is linked not only to memory retrieval after stress - as noted earlier - but also to the formation of new memories (42). P05067, amyloid-beta precursor protein (APP). APP is a cell surface receptor that performs physiological functions related to neurite outgrowth, neuronal adhesion, and axonogenesis on the neuronal surface. The interaction of APP molecules on neighboring cells promotes the formation of synapses and amyloid fibrils. App were associated with memory impairment, and APP may contribute to the cognitive deficits associated with Alzheimer’s disease.(43).

P13591, Neural cell adhesion molecule 1 (NCAM1). NCAM1 is involved in several brain-related biological processes, including neuronal migration, axon branching, fascicles, and synaptogenesis, and plays a key role in synaptic plasticity. (44). Stress and glucocorticoid signaling transiently activate NCAM expression in memory-associated brain regions to promote long-term memory formation(45). NCAM1 is involved in the acquisition and expression of emotional memory (46). P01019 and angiotensinogen (AGT) are involved in many biological processes, including cell‒cell signaling and regulation of long-term neuronal synaptic plasticity (www.uniprot, org). P15289 and Arylsulfatase A (ARSA) have been reported to impair memory in ASA knockout mice. (47).

All patients had symptoms of memory impairment to different degrees after MECT. The above results showed that the common proteins were CSPG4, CBG, APP, NCAM1, and ARSA, which are related to memory loss, memory damage and memory formation.

The connection between proteins and pathways of MECT reflected in urine is shown in Fig. 6. The types of pathways mainly included cell adhesion, immune response, axon guidance, and oxidation stress. These pathways and proteins induce neurogenesis and plastic synapses, consistent with the MECT treatment mechanism.

**Fig. 6.**
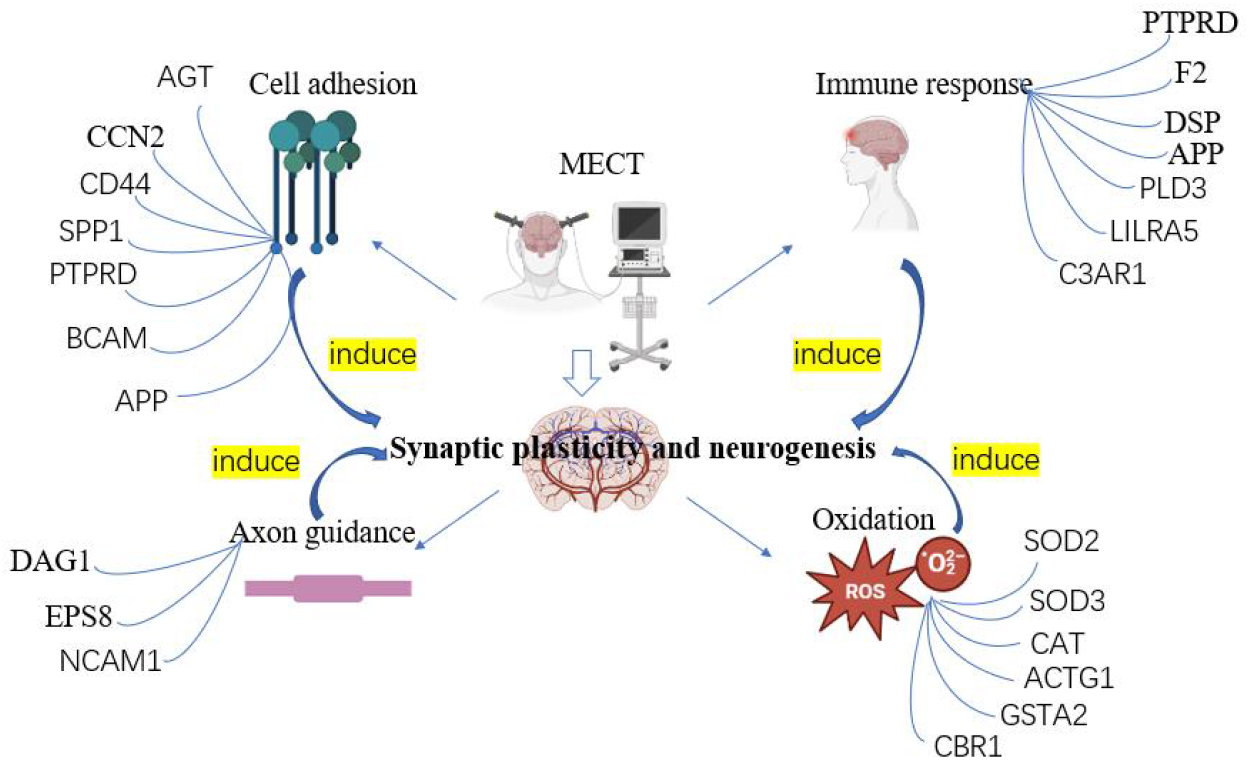
Differential proteins and biological processes related to the MECT mechanism

The present study is the first clinical study to explore the antidepressant mechanism of MECT through urinary proteome profiling. In this study, self-control was performed for patients with remission of depressive symptoms to exclude the influence of confounding factors (drugs, age, heterogeneity of depression, etc.), to ensure that common differential proteins and functions shared by most patients were most likely to indicate changes in MECT in urine. The high complexity of depression makes self-control an important method for future proteomic studies of depression. MECT is also effective for other psychiatric diseases in addition to depression, so neuroplasticity may be related to the treatment of other psychiatric diseases. The sample size of this study was insufficient. In the future, it is necessary to control the MDD sample subtypes and increase the sample size to verify the effect of MECT.

## 4 Conclusion

In summary, we attempted to explore the urinary manifestation of brain activity-related changes after MECT. Our results indicated that the treatment of depression by MECT was individual. Common biological processes and common proteins were closely related to neurogenesis and synaptic plasticity, which is consistent with the mechanism of MECT. Our study provides clues for the mechanistic exploration of MECT in the treatment of MDD.

## Data Availability

All data produced in the present work are contained in the manuscript

## Acknowledgements

We are especially grateful to professor Gang Wang’ s group of Depression Treatment Center, Beijing Anding Hospital, Capital Medical University for sample collection and providing ethical review. This work was supported by grants from STI2030-Major Projects(No. 2021ZD0200600)and Beijing Hospitals Authority Youth Programme(No. QML20211902).

